# A framework for making predictive models useful in practice

**DOI:** 10.1101/2020.07.10.20149419

**Authors:** Kenneth Jung, Sehj Kashyap, Anand Avati, Stephanie Harman, Heather Shaw, Ron Li, Margaret Smith, Kenny Shum, Jacob Javitz, Yohan Vetteth, Tina Seto, Steven C. Bagley, Nigam H. Shah

## Abstract

**Objective:** To analyze the impact of factors in healthcare delivery on the net benefit of triggering an Advanced Care Planning (ACP) workflow based on predictions of 12-month mortality.

**Materials and Methods:** We built a predictive model of 12-month mortality using electronic health record data and evaluated the impact of healthcare delivery factors on the net benefit of triggering an ACP workflow based on the models’ predictions. Factors included non-clinical reasons that make ACP inappropriate, limited capacity for ACP, inability to follow up due to patient discharge, and availability of an outpatient workflow to follow up on missed cases. We also quantified the relative benefits of increasing capacity for inpatient ACP versus outpatient ACP.

**Results:** Work capacity constraints and discharge timing can significantly reduce the net benefit of triggering the ACP workflow based on a model’s predictions. However, the reduction can be mitigated by creating an outpatient ACP workflow. Given limited resources to either add capacity for inpatient ACP versus developing outpatient ACP capability, the latter is likely to provide more benefit to patient care.

**Discussion:** The benefit of using a predictive model for identifying patients for interventions is highly dependent on the capacity to execute the workflow triggered by the model. We provide a framework for quantifying the impact of healthcare delivery factors and work capacity constraints on achieved benefit.

**Conclusion:** An analysis of the sensitivity of the net benefit realized by a predictive model triggered clinical workflow to various healthcare delivery factors is necessary for making predictive models useful in practice.

## Introduction

Over the past decade, the rapid increase in the availability of healthcare data collected during routine care and dramatic advances in machine learning have fed a great deal of excitement about using machine learning to improve clinical care ^1–4^. Predictive models, which estimate the probability of some event of interest occuring in a specified time frame in the future, have been developed for events such as heart failure, inpatient mortality, and patient deterioration ^5^. However, there have been relatively few success stories where these models led to impact on what matters to patients, providers, and healthcare decision makers such as, reduction in costs, lower rate of those clinical events, and increased access to care ^6,7^.

The lack of impact is in part because the evaluation of machine learning models typically focuses on measures of performance, such as area under the receiver-operator curve (AUROC) and mean precision. Such measures provide a convenient quantitative summary of the performance of the model, but they do not reflect the consequences of taking action based on the model’s output ^8^. The translation of model performance into benefit to patient care is mediated by multiple factors related to existing and proposed clinical workflows that incur a broad range of data acquisition, change management, and workflow integration burdens ^9–11^. While measures such as AUROC can summarize the accuracy of predicting an event, they have little bearing on the effectiveness of actions taken in response to the prediction, the constraints on those actions ^8^, and the ethical concerns or value mismatches that may arise from those actions ^12–14^. In many cases the overall benefit of using predictive models for improving healthcare will be critically determined by the considerations of implementation costs, actionability, safety and utility ^6,9,11^.

In this work, we present a comprehensive analysis of the net benefit of a machine-learning enabled workflow for identifying patients for Advance Care Planning (ACP). ACP is the elicitation and documentation of patient values and preferences regarding goals of care. Documentation of ACP is critical for guiding care when patients are seriously ill and unable to express their wishes or make their own decisions. The target population is patients admitted to the General Medicine service at Stanford Hospital. The prediction is an estimate of 12-month mortality based on the patient’s historical electronic health record (EHR) data; the action triggered by a high-risk estimate is the offering of ACP, carried out by the General Medicine teams. Timely offering of ACP and palliative care have proven to be effective at both reducing downstream costs ^15–17^, improving physician morale, and in some cases, improving outcomes such as survival ^18,19^. However, despite widespread recognition that such interventions are underutilized ^20–24^, timely ACP by hospitalists can be difficult, given the urgency of taking care of a severely ill patient, which can interfere with consideration of a patient’s longer term prognosis.

Predictive models for mortality have been evaluated for identifying patients to receive ACP or similar interventions ^25–28^. There is evidence that care workflows using such predictive models to screen patients can identify the right patients for timely conversations about goals of care ^27^. However, over the course of designing an implementation of a workflow enabled by a machine learning model at Stanford Hospital in early 2020, we identified various factors that limit the presumed net benefit resulting from setting up a model-triggered workflow. First, there may be limited capacity to perform ACP even if patients are correctly identified by the model. Second, effective ACP takes time to conduct since it is based on conversations with patients and families. Building the shared understanding and rapport necessary for these conversations takes time which may be difficult to find for busy physicians. Third, it is possible to imagine alternatives that can mitigate the impact of these factors, such as increasing our capacity for inpatient ACP by hiring more staff, or developing the capability for outpatient ACP. We quantify the impact of these factors and gain insight into how best to mitigate those impacts.

Our approach is inspired by the evaluation of screening tests, and we view the prediction model as a screening test that triggers follow-up care. For example, analogous to a number needed to screen, we can compute a number needed to benefit which calculates the number of patients that would need to be screened by a test and treated in response to a positive result ^29^. Cost-benefit analysis is a widely used approach to evaluate decisions made under such uncertainty^30^; here we apply a cost-benefit analysis on multiple factors impacting the effectiveness of the proposed workflow using simulations and sensitivity analysis.

The essence of our approach is to estimate utilities for each of four possible outcomes derived from the model output for each patient: true positives (the model correctly flags a patient for ACP and ACP is carried out), false positives (the model incorrectly flags a patient for ACP and it is carried out), true negatives (the model correctly fails to flag a patient for ACP and none is carried out), and false negatives (the model incorrectly fails to flag a patient for ACP, and no ACP is carried out). The utility for each outcome quantifies the benefit associated with that outcome, in whatever units are most convenient or meaningful. In this work, we quantify utility as *total healthcare expenditures in the six months following discharge* because estimates are available from a multi-site RCT ^15,31^. If utility estimates are available in standard units of the amount of unwanted care avoided, or increase in patient comfort, the framework would still apply.

Given these RCT derived utilities, along with a set of patients for whom we have both risk estimates and expert-provided, ground-truth assessments of appropriateness for ACP, we can then estimate the benefit under the setup of screening by a model, and compare it to the benefit achieved by policies such as intervening on all or none of the patients. While such approaches have been advocated for evaluating predictive analytics in healthcare ^9,32,33^, they likely overestimate the benefit realized in practice, because factors such as limited capacity to perform ACP can reduce the effective number of true positives. For example, if we can only perform ACP for one patient per day, then no benefit is accrued by the second and third patients flagged for ACP even if the predictions are correct. In this study, we quantify the effect of such healthcare delivery factors by performing simulations that assess a broad range of failure rates for a variety of factors. We also assess an alternative workflow in which ACP is conducted via an outpatient pathway for patients who were correctly flagged for ACP by the model, but for whom ACP was not completed during their hospital stay.

## Methods

Our analysis uses simulations of a proposed prediction triggered workflow (i.e., the responsive *action*) running for an extended period. The prediction-action dyad starts with an automated estimation of the risk of 12-month all-cause mortality for patients admitted to the General Medicine service at Stanford Hospital. Risk estimates are output by a model for all-cause mortality, as described below. Chart review by an experienced palliative care nurse was used to assign ground-truth labels for whether ACP was appropriate. Patients whose risk estimate exceeds a chosen threshold are considered referred for an ACP intervention. Given a risk threshold, we can calculate a utility achieved for each patient, using the risk estimate and the ground-truth label to categorize that prediction as true positive, false positive, true negative, or false negative. Simulations vary the effects of various factors, such as work capacity constraints, which may prevent ACP from being successfully carried out, thus reducing the utility actually achieved for the patient. Below, we describe the predictive model, its validation via chart review, and our simulation setup.

### Model development

We developed a gradient boosted tree model ^34^ to estimate the probability of 1-year all-cause mortality upon inpatient admission, using a de-identified, retrospective dataset of EHR records for adult patients seen at Stanford Hospital between 2010 and 2017 ^35^. This dataset comprised 97,683 admissions, and had a prevalence of 17.6% for one-year all-cause mortality. Data were obtained under a Stanford University Institutional Review Board approved protocol, and informed consent was waived. The input data for a given admission consisted of basic demographic information (age and gender), along with counts of diagnosis codes, medication orders and encounter types (e.g., inpatient, office visits, surgery) observed in the year prior to admission. A total of 63,043 features were used. We split the admissions according to year: we used 82,525 admissions from 2010 through 2015 as training data, and 15,098 admissions in 2016 and 2017 as validation data. We tuned hyperparameters based on performance in the validation data, and trained a final model on the full dataset using the optimal hyperparameters. We evaluated the performance of the resulting model against clinician chart review prospectively, as described below. We omitted admissions for which we do not have reliable indications of survival (e.g., in person encounters) or mortality in the following year. Although our analysis in this work is focussed on admissions to the General Medicine service, the 1-year all-cause mortality model was developed using admissions to all services. We did so to ensure adequate training data and to obtain a prediction model that can be used for other service lines as well.

### Prospective evaluation by chart review

We conducted a prospective evaluation of the model against expert chart review over 2 months in the first half of 2019; each day in that period we presented an experienced palliative-care advanced-practice (AP) nurse with a list of patients newly admitted to General Medicine at Stanford Hospital. These lists were in random order, and no model output was presented. These data were collected under a separate Stanford University Institutional Review Board approved protocol; need for informed consent was waived. The AP nurse performed chart reviews for each patient in order, as time permitted each day, to answer the question, “*Would you be surprised if this patient passed away in the next twelve months?*” On any given day, the evaluator might not have been able to complete the chart review for all, or even any, of the newly admitted patients. However, the set of patients for whom chart review was completed is random. We used the responses as a ground-truth label for appropriateness for ACP because in the current, sans model setup, such assessment is a key determinant for triggering a consultation with palliative care. Of the 191 patient charts reviewed, we were able to map 178 to the de-identified EHR data extract used for model development. The median age was 61 years (minimum 21 and maximum 88), and 48.5% were male. The AP nurse responded “No” to the surprise question for 29.2% of these patients. This population had 21.9% patients presenting with infection, 13.5% with gastrointestinal problems, and 12.9% with cancer. The model achieved an AUROC of 0.86 (95% confidence interval of 0.8-0.92, estimated from 1000 bootstrap samples of the chart review set).

### Utility values and simulation parameters

We estimated the utilities of the four possible outcomes of an intervention, U_tp_, U_fn_, U_fp_, and U_tn_, using a multi-center randomized trial of inpatient palliative care consults ^15^ to measure the post-discharge health care costs (Table 1). Gade et al. found that patients who received usual care incurred an average cost of $21,252 in the six months following discharge, while patients who received an inpatient palliative care consultation incurred an average cost of $14,486. The consultation itself cost $1,911 for a net savings of $4,855. These data were collected between 2002 and 2003; accounting for inflation ^36^ yields costs of $28,613 and $37,085 for patients who do and do not receive ACP respectively. We used these estimates as the values for true positives and false negatives, respectively. We estimated the cost of true negatives and false positives as follows: For true negatives, we used half of the mean per capita annual spending for patients in “poor” health determined from the Peterson-Kaiser Health System Tracker ^37^ because this population is similar to General Medicine patients at Stanford Hospital. For false positives, we added the cost of the intervention to this cost, yielding a total of $14,970. These utility values are summarized in Table 1.

**Table 1.**
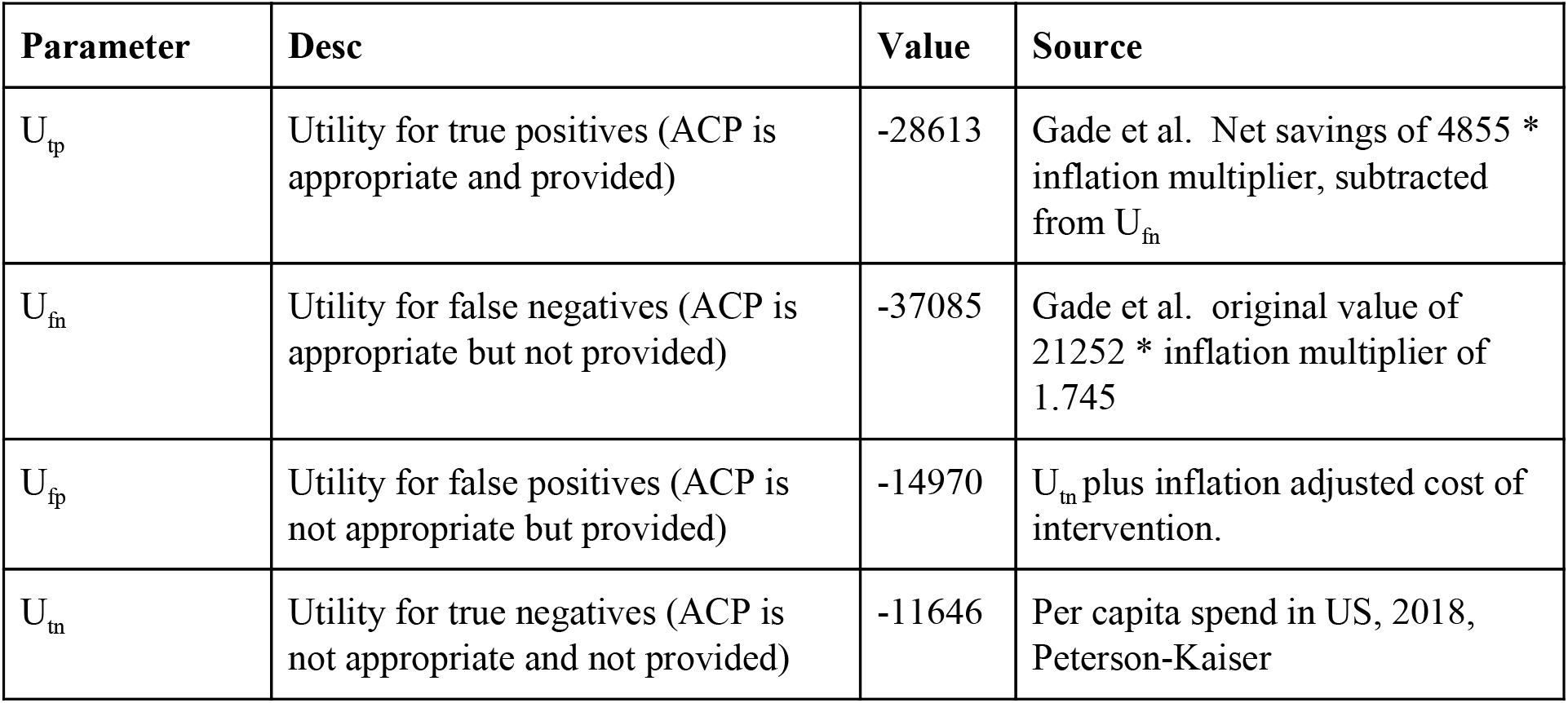
Utility values

### Establishing the “best case”

Our simulation relies on resampling of the 178 admissions for which we have clinician evaluations, model output, and length of stay. To establish the “best case”, we simulated 5000 days of offering ACP. Each simulated day, we picked the number of admissions from the empirical distribution of admissions per day observed from March through August 2019 (the mean number of admissions to the General Medicine service was 9.4 per day, with a standard deviation of 3.4). We then chose that many admissions with replacement from the 178 admissions with model output and chart review. We calculated the number of patients in each of the four possible outcomes at each possible threshold of the model output to get the true positives, false negatives, false positives, and true negatives for the day. These counts were multiplied by the utilities for each category (U_tp_, U_fn_, U_fp_, and U_tn_), and the products summed. The result is the utility achieved on a given day, which is divided by the number of admissions to yield a per-admission utility.

We simulated 5000 such days, and calculated the mean utility over simulated days at each possible threshold. We smoothed the mean utilities across thresholds using loess with a smoothing parameter of 0.75 ^38^, and took the maximum utility across all possible thresholds. We then normalize this maximum to the status quo ante by subtracting the utility under a policy of Treat Nobody, yielding the *possible maximum net utility*. Note that establishing the “best case” in this manner assumes each true positive is translated into successful execution of the ACP workflow.

### Impact of external factors

Our next objective is to explore how various factors impact the achieved utility using the best case as a reference. The factors explored are listed in Table 2.

**Table 2.** Simulation parameters to explore the impact of external factors

| Parameter | Desc | Value | Range |
| --- | --- | --- | --- |
| Rejection rate | Fraction of patients for whom ACP is not possible for non-clinical reasons | 0.1 | 0.1, 0.2, 0.3 |
| Daily capacity | Daily capacity to carry out ACP | 3 | 1, 2, 3, 4, 5 |
| Mean time to complete ACP | Mean time in days to complete ACP, parameter to exponential distribution. | 2 | 1, 2, 3, 4 |
| Outpatient pathway rescue rate | Rate of successful ACP by outpatient pathway. | NA | 0, 0.25, 0.50, 0.75, 1.0 |

### Rejection for non-clinical reasons

Often, a patient recommended by a model is a valid candidate for ACP with respect to clinical prognosis, but has declined prior attempts at goals-of-care conversations. We simulated this factor by flipping a weighted coin for each true positive patient in the best case simulation. The weighted coin represents a Bernoulli trial with some probability that ACP will be rejected for non-clinical reasons. We varied the probability of rejection as 0.1, 0.2, and 0.3. We normalized the maximum utility achieved across all possible thresholds against the utility of the status quo ante to yield a net utility as before, and compared against the net utility of the best case. We present all results as a fraction of the best case utility to avoid tying the analysis with specific dollar values, and ensuring that if utility estimates are available in standard units of the amount of unwanted care avoided, or increased in patient comfort, the approach can still be used.

### Limited capacity for ACP

Another frequent situation is that a patient is a valid candidate but the clinician has limited time during the day to initiate ACP; i.e. the “crazy day in the clinic”. We simulated this factor as a hard constraint on the capacity of the General Medicine service to intervene. We proceeded similarly to the best case simulation except that for each day we capped the number of true positives to some fixed limit (1, 2, 3, or 4). Any true positives in excess of this limit were counted as false negatives. As before we compared the net maximum utility under each scenario against that of the best case.

### Failure to complete ACP due to early discharge

A significant fraction (57%) of hospital stays are three days or less, while ACP may take several days due to logistical constraints, such as scheduling time with family members. For each true positive patient, we sampled the time required to complete ACP from an exponential distribution with different means; representing scenarios in which it takes 1, 2, 3, or 4 days on average to complete ACP. If the sampled time to completion exceeds the true length of stay for the patient, we count the patient as false negative from failure to complete ACP due to early discharge. As before we compared the net maximum utility under each scenario against that of the best case.

### Effect of an outpatient ACP pathway

An outpatient care pathway for ACP may significantly mitigate decreases in achieved benefit from factors such as capacity constraints or failure to complete ACP due to discharge. To simulate this, we flipped a weighted coin for each patient who was moved from the true positive to the false negative categories due to a capacity constraint or early discharge. This coin flip represents the probability that the outpatient ACP pathway will be successful in serving the patient. We varied this probability from 0, 0.5 and 1.0, while keeping rejection for non-clinical reasons at 10%, a capacity constraint of three ACP interventions daily, and a mean time to completion of two days. As before, we compared the net maximum utility under each scenario against that of the best case.

### Trade offs between inpatient capacity vs outpatient ACP

Of the failure modes analyzed, we have the least control over rejection for non-clinical reasons. Similarly, it is not reasonable to extend the length of stay solely to complete ACP, especially given that length of stay is a closely watched quality metric. Therefore, the two factors that are most amenable to modification are capacity to offer ACP in the inpatient setting, and serving patients via an outpatient ACP pathway.

We therefore explored the relative efficiency of increasing inpatient capacity versus improving “rescue” via an outpatient pathway. We simulated 5000 days at each combination of a range of inpatient capacities (1 through 5) and rescue rates (0%, 50%, and 100%). We then calculated the net maximum utility under each scenario and compared the net maximum utility gained by increasing inpatient capacity by one versus improving the outpatient pathway.

### Analysis of utility ranges and effects of work capacity limits

Our simulation analysis used fixed utility values for the true positives, false negatives, false positives, and true negatives. These values might differ at different sites, and in some cases it may not be possible to obtain exact values. Therefore, to analyse the effect of choosing different utilities, each of the four entries in a utility matrix were expanded symmetrically by 10% to form the lower and upper bounds of *utility ranges*. For each combination of lower and upper bound of all four ranges, we computed the maximum expected utility for acting on the model’s predictions, producing 2^4, or 16, possible maxima.

Separately, we constructed a graphical representation showing the relationship between choosing different classifier probability thresholds, the expected utility, and the total number of predicted positives, both true as well as false. Each positive prediction is a patient who will be offered an ACP consult. We refer to the total number of positives on whom to follow up as “work”.

## Results

We performed a series of simulation studies to evaluate the impact of multiple factors on the net utility of a workflow for ACP triggered by a 12-month mortality prediction. The factors analyzed were: (1) ineligibility of candidates due to non-clinical reasons, (2) limited capacity to offer ACP, (3) failure to complete ACP due to early discharge, and (4) availability of an outpatient pathway to follow up on missed cases. We also evaluated the relative benefits of increasing capacity for outpatient ACP versus increasing inpatient ACP capacity. We found that work capacity constraints and failure to complete ACP due to early discharge can significantly reduce the benefit. The impact of these factors can be mitigated by an alternative pathway for offering ACP in the outpatient setting. Under resource limitations to either add capacity for conducting inpatient ACP versus developing the outpatient pathway, the latter is likely to provide more benefit to patients.

### Impact of external factors on the realized benefit

#### Rejection for non-clinical reasons

Figure 1a shows the fraction of the best case, per patient utility achieved, as the rate of rejection for non-clinical reasons varies from 0% to 30%. We see a decrease in the per patient utility at all thresholds. In the best case scenario (no rejections), we would have seen an average utility of -$17,701 per patient. This best case sets the maximum achievable benefit and the ‘not offering ACP’ sets the zero baseline in the y-axis of the figure. At the 10% rejection rate scenario, we achieve the majority of the ‘best case’ utility, with a linear decline as more patients are rejected for non-clinical reasons. At a 30% rejection rate, we achieve just short of 75% of the best case. We present all results as a fraction of the best case utility to avoid tying the analysis with specific dollar values, and ensuring that if utility estimates are available in standard units of the amount of unwanted care avoided, or increased in patient comfort, the approach can still be used.

**Figure 1.**
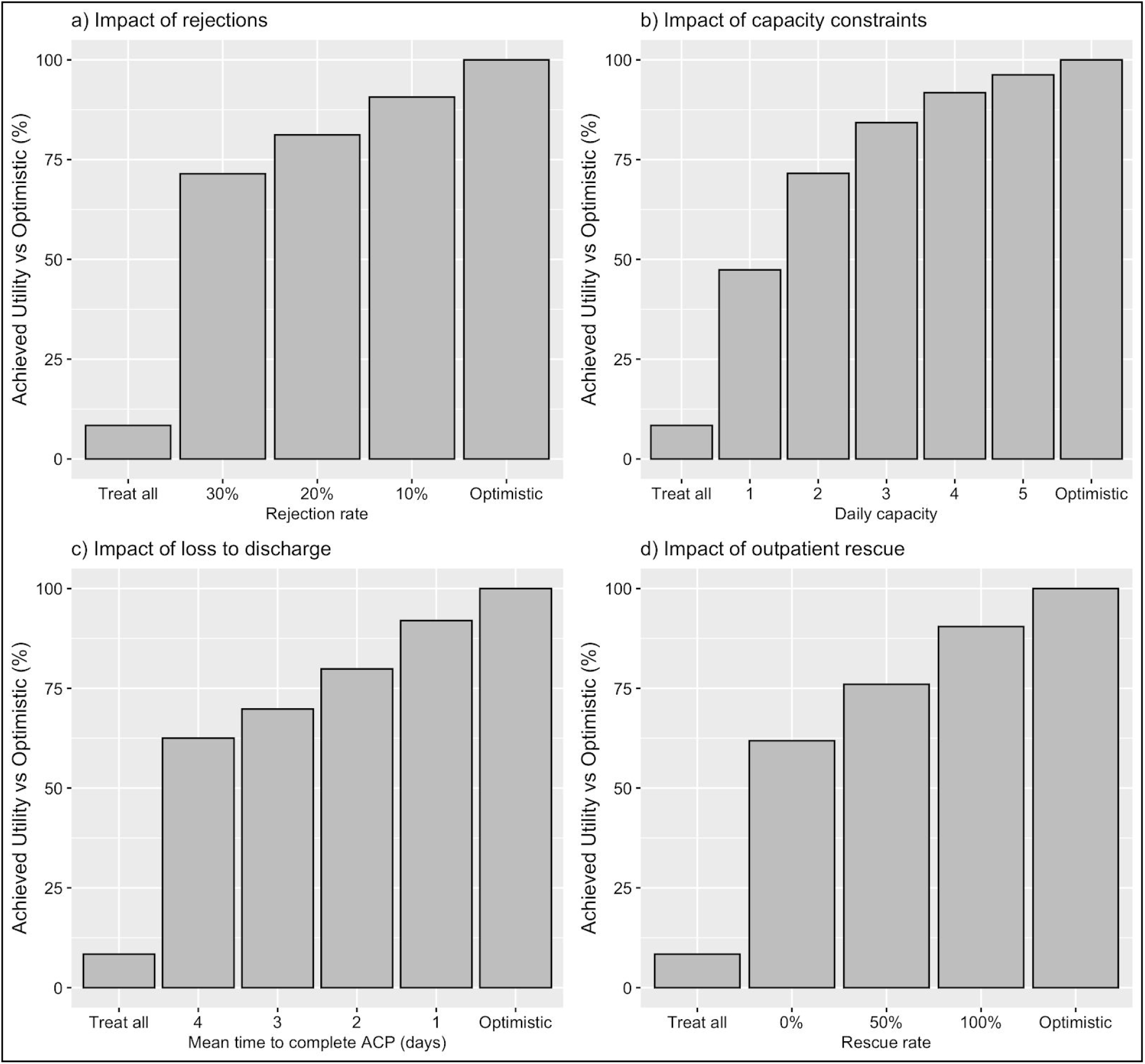
The figure summarizes the effect of different factors on the realized net utility of triggering a care workflow based on a predictive model for one year mortality. In all plots the y-axis shows the achieved net utility relative to the best case labeled as ‘optimistic’. The default state of treating nobody, is the 0 point on the y-axis. The achieved utility is plotted as a percentage of the best case scenario, in which every prediction is followed up by ACP. We also plot the relative net utility of treating everybody (Treat all) for comparison. **A. Impact of rejection of recommendations for ACP for non-clinical reasons**. The x-axis shows the rate of rejection of ACP due to non-clinical factors ranging from 10% to 30%. The rejection rate translates to a linear reduction in net utility. **B. Impact of capacity constraints on per patient utility**. The x-axis shows different capacity constraints for conducting ACP. Capacity constraints have a large impact on net utility, with a capacity of 1 capturing close to 50% utility of the “best case”. Increasing capacity offers rapidly diminishing returns because there are fewdays when more than four patients are recommended for ACP. **C. Impact of failure to complete ACP due to discharge on per patient utility**. The x-axis shows the average number of days it takes to complete ACP. The relative net benefit ranges from 92% to 62.5% of the best case estimate as the mean time to complete ACP ranges from one to four days. **D. Impact of an outpatient rescue pathway on per patient utility**. The x-axis shows the effect of rescuing 0%, 50%, and 100% of the model’s recommendations. Without rescue, the net utility is 65% of the optimistic estimate. At 50% rescue, we achieve 76% of the optimistic estimate. At 100% rescue, we achieve 90.5% of the best case scenario because the outpatient rescue pathway can not rescue ACP rejected for non-clinical reasons.

#### Limited capacity for ACP

The General Medicine service takes care of complex patients and clinicians often do not have time to perform ACP. As described in the methods, we simulated the impact of this as a limit on the number of patients for whom ACP can be done each day. The results are shown in Figure 1b. We see that having a work capacity of just one ACP per day captures just shy of half of the best case achieved benefit. Increasing the ACP capacity yields rapidly diminishing returns, with very little difference at a capacity of 5 compared to the “best case” scenario. This is because there are very few days in which we can expect to need ACP for four or more patients.

#### Failure to complete ACP due to early discharge

Because the ACP intervention takes time to perform, clinicians may not be able to complete ACP before the patient is discharged. We simulated this loss to discharge as a random process as described in the methods. Figure 1c shows that failure to complete ACP due to discharge may have a significant impact on achieved utility. If it takes on average two days to complete ACP, the savings relative to the best case drop to about 75%. If it takes four days on average to complete ACP, the impact on net benefit drops to 62.5% of best case.

#### Effect of an outpatient ACP Pathway

Based on our results, capacity constraints and failure to complete ACP due to discharge could significantly reduce the benefit of a prediction triggered ACP workflow. Therefore, we explored the potential impact of having a pathway for performing ACP in an outpatient setting, post-discharge. We fixed a 10% rate of rejection due to external factors, along with a capacity constraint of three ACP per day, and a mean time to complete ACP of two days. We vary the rate at which the outpatient pathway successfully completes the recommended ACP from 0%, 50% and 75%. Figure 1d shows the impact of the proposed outpatient pathway. Without outpatient rescue, the maximum achievable benefit is 66% of the best case, which is significantly down from the near-best-case seen with a 100% rescue rate. However, even a 50% rescue rate recovers a significant fraction of the best case net benefit.

### Trade offs between inpatient capacity vs outpatient ACP pathway

Of the factors we examine in this study, the modifiable ones are capacity for inpatient ACP and creation of an outpatient ACP pathway. The relative benefit of these alternatives matter because they are likely to have significantly different set up and operating costs. We examine the relative benefit of increasing inpatient ACP capacity by one patient, versus investing in an outpatient pathway for ACP, at various starting levels of inpatient ACP capacity.

Figure 2 shows the change in mean per patient utility as we increase inpatient capacity starting with different initial inpatient capacities (solid red line). The figure also shows the change in mean patient utility with an outpatient pathway for ACP with 25% to 100% success rates. We find that no matter what the starting inpatient capacity is, an outpatient pathway with a 50% success rate (dashed green line) results in greater savings than increasing inpatient ACP capacity.

**Figure 2.**
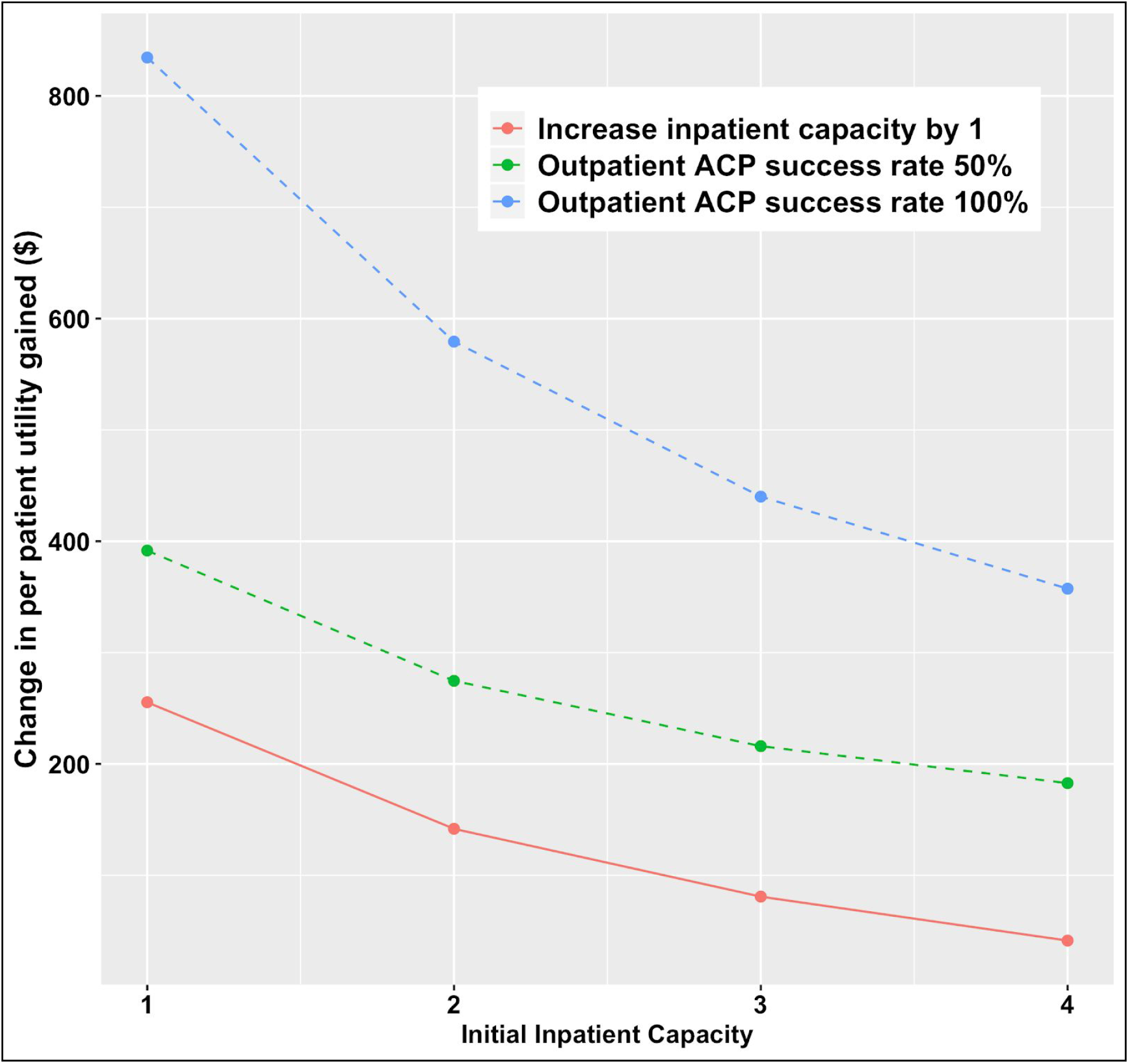
Tradeoff between adding inpatient capacity for ACP versus outpatient capacity. The plot shows the change in mean per patient utility as we increment inpatient capacity starting from different initial inpatient capacity (solid red line). The dashed lines show the change in mean patient utility for having an outpatient pathway for ACP with 50% and 100% success rates. We find that at all starting inpatient capacities, an outpatient pathway with even a 50% success rate results in greater utility than adding to inpatient capacity.

### Analysis of utility ranges and effects of work capacity limits

The analysis thus far used fixed utility values, which can be hard to obtain or simply differ by site. Therefore, as described in the methods, each of the four entries in a utility matrix were expanded by +/- 10% to form utility ranges. Examining all possible combinations of utility ranges gives a range of -$15800 to -$19300 for the maximum expected per-patient utility.

Whatever the value of maximum expected per-patient utility may be, there exists a tradeoff between per-patient utility and work capacity as illustrated in Figure 3. The x-axis is the probability threshold on the output from the classifier, ranging from 1.0 down to 0.0. Depending on the chosen threshold, patients with a predicted probability higher than the threshold will be followed up on. The y-axis is the expected per-patient utility calculated using the four utility values from Table 1. The boxed numbers -- located at regularly-spaced probability thresholds -- are the patients to follow up on (i.e. work), expressed as a percentage of the total patients for whom a prediction is made.

**Figure 3.**
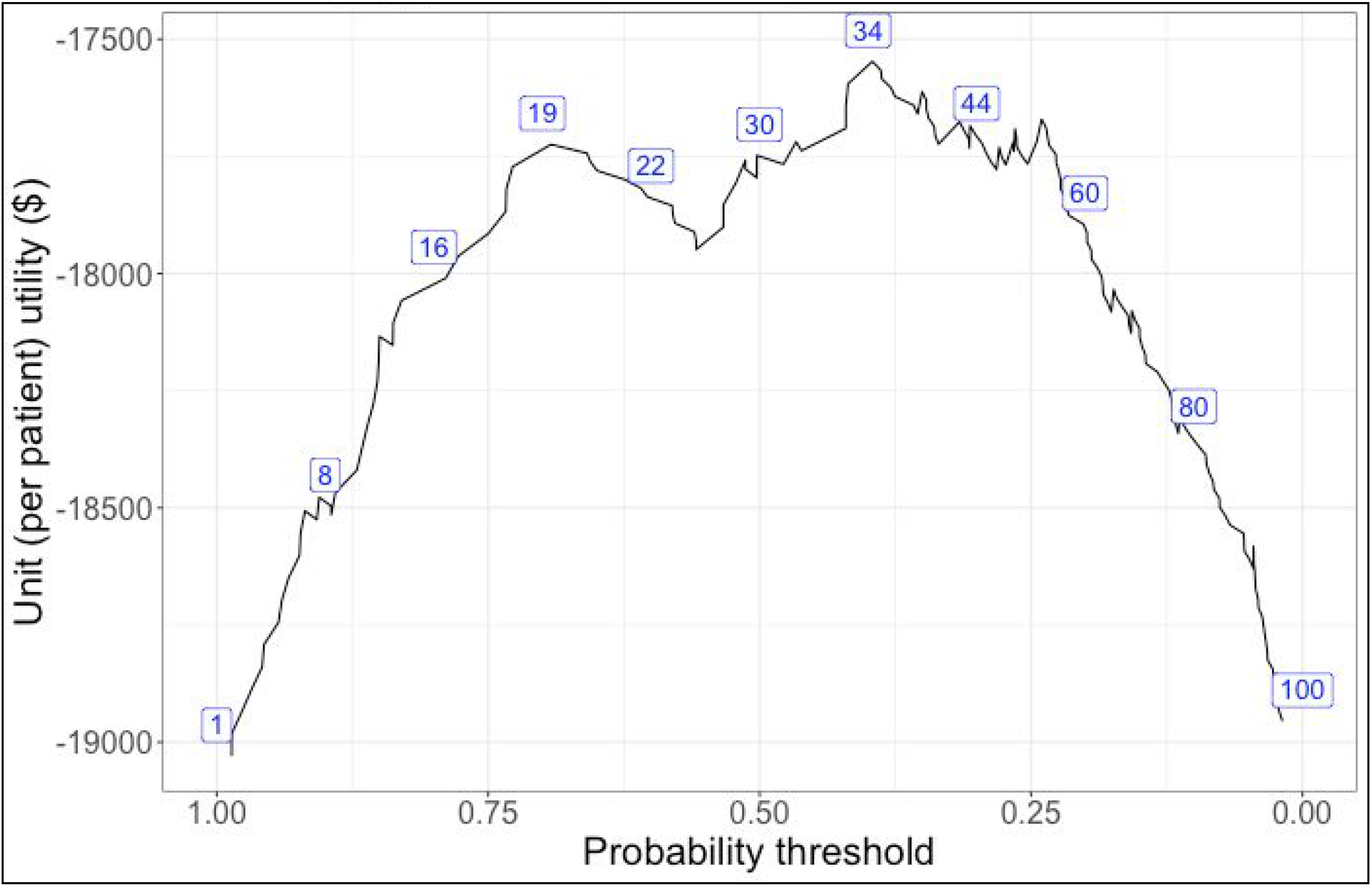
Unit (per-patient) utility versus the probability threshold at which a patient is referred for follow up. The boxed numbers are the number of patients to follow up with (true positive and false positive), or “work” at that threshold, expressed as a percentage. Work increases as more patients are referred for ACP consultation. There is a tension between the goal of maximizing total utility, which is the product of per-patient utility and the number of patients acted upon; while keeping the number of patients followed up below the hospital system’s work capacity limit.

For our model, the number of new cases for which predictions would be made corresponds to the new admissions per day to the Gen Med service (mean 9.4 patients, std. dev. 3.4). In this situation, a work capacity of 3 (which is 23.4 % of the mean + one std. dev. of daily admits) is not close to the maximum per-patient utility, reinforcing the need to consider alternatives, such as an outpatient ACP pathway.

## Discussion

Despite considerable improvements in measures of performance of predictive models in healthcare, there have been relatively few successes using such models to provide better clinical outcomes at lower costs ^39,40^. We believe that this is in part because the translation of modeling advances into improvements in clinical care requires integrating the model’s output into complex human workflows that are separate from model performance ^6,8,9^. If the actions taken in response to a predictive model are embedded in such a system, the ability to make predictions, and improvements in predictive accuracy alone, are not sufficient to improve care.

We note that successful uses of ML in healthcare typically require considerable integration effort after model development is done. For example, the majority of published efforts regarding the sepsis early warning system (EWS) deployed at Kaiser Permanente Northern California focus on understanding existing processes and then managing the iterative refinement and dissemination of new workflows supported by the EWS model ^41–44^. This experience reinforces the lesson learned from decades of quality improvement efforts: clinical care takes place in a complex, adaptive social environment, and changing processes within that environment is not amenable to purely technical interventions.

In this study of a predictive model for mortality used to recommend patients for ACP, we identified several workflow factors that limit the benefit of setting up a model-triggered care workflow, such as limited capacity for ACP during inpatient admissions, and early discharge before ACP can be completed. These factors partially explain the critiques that models are proliferating but concrete benefit is scant ^6,39^. Our analyses used simulations examining the net-utility of workflows triggered by the model to elucidate the impact of workflow factors. These analysis techniques are not new, but have not received sufficient attention in the ML-for-healthcare community ^33,45,46^. We believe it is time to adopt methods from health delivery science and health services research to provide honest evaluations of machine learning guided interventions in healthcare ^47^ and to develop a delivery science for AI interventions in healthcare^48.^

It is natural to ask where do such analyses fit into the development and deployment path for predictive models into the clinic. We adopt an idealized four-stage framework, presented in Figure 4, for such projects. The first stage focuses on clear definitions of both the modeling problem (i.e., what is the prediction target, how is it defined, when does the prediction happen, and what data are available at that time) and, just as important, the intervention presumably triggered by the model’s output, along with a clear articulation of the desired benefits sought. The second stage consists of development and technical validation of the prediction model; this is the focus of the bulk of currently published work in the machine learning for health community ^40,49,50^. The third stage comprises careful, iterative development of the clinical workflow associated with the model: what are the set of actions to be undertaken in response to a prediction? The final stage consists of monitoring and maintenance of a model and associated workflows, ideally followed by a prospective trial to demonstrate efficacy.

**Figure 4.**
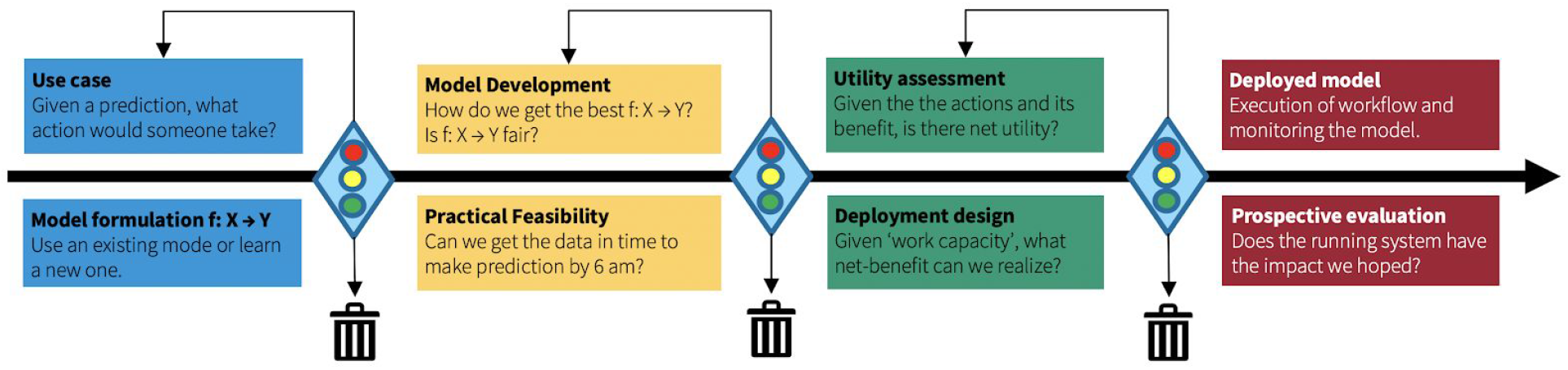
A four-stage framework guiding the development and evaluation of a predictive model throughout its life cycle. The stages are: 1) problem specification and clarification, 2) development and validation of the model, 3) analysis of utility and impacts on the clinical workflow that is triggered by the model, and 4) monitoring and maintenance of the deployed model as well as evaluation of the running system comprised of the model-triggered workflow.

Analyses such as the one presented here are for guiding stage 3 (green) of the process in figure 4. In this stage, the model itself takes a backseat, and we focus on where it fits into the clinical workflow, and how to alter that workflow to achieve the desired benefit. Such analyses, when used in conjunction with traditional methodologies for change management and implementation science, can provide valuable guidance on where to focus efforts and resources in order to close the gap between best case estimates of benefit and the realized benefit.

Such analyses have limitations. First and foremost is that it can be very difficult to estimate true cost and thus calculate utilities ^45,46^. In this work, we have used post-discharge healthcare expenditures as a stand-in for utility; however, we acknowledge that this is a convenience view that allows us to illustrate the need for such analysis in the absence of a standard quantification mechanism for the amount of unwanted care avoided, or increased in patient comfort. We note that the ultimate purpose of ACP is to provide care that is concordant with the expressed values and wishes of patients, that this provides benefits for providers who seek to provide the best care possible for their patients, and that this “true goal” is not easily measured and not captured by downstream healthcare spending. Therefore cost as a measure, while limited, is still directionally useful and we present results as a fraction of the best case utility to avoid tying the analysis with specific dollar values. A second significant limitation of this study is that we used an expert’s response to the question, “Would you be surprised if this patient passed away in the next 12 months” as a source for our ground-truth labels. This mirrors one of the key criteria that is supposed to trigger ACP in current workflows, but it is possible that an expert may be incorrect in their judgement. Finally, our simulations are not sufficiently fine-grained to accurately capture the subtleties of all the factors in play. These limitations notwithstanding, we believe that the analyses presented is a useful template for how to obtain insight into the net effect of a proposed predictive model paired with a clinical intervention.

## Conclusion

We analyzed the impact of factors in healthcare delivery on the realized benefit of using a predictive model for 12-month mortality to identify patients for ACP. The analyses use simulations to identify factors that have a large impact on the achieved benefit of using the model to trigger an intervention. Factors included non-clinical reasons that make ACP inappropriate, limited capacity for ACP, inability to follow up due to patient discharge, and availability of an outpatient workflow to follow up on missed cases. The resulting estimates of the impact of these factors can guide allocation of resources to mitigate reductions in achieved benefit. We argue that routine use of such analyses of the sensitivity of the net benefit to various healthcare delivery factors is necessary for translation of advances in predictive modeling into real world clinical benefit.

## Data Availability

The data are patient data, and hence not available publicly.

## Acknowledgements

The study was supported by the Stanford Medicine Program for AI in Healthcare, which is enabled by the Department of Medicine, a research grant from Google, an endowment from Debra and Mark Leslie, and innovations funds from Stanford Healthcare. We acknowledge Ethan Steinberg for providing feedback on the analysis of work capacity limits and helpful commentary by Dr. Birju Patel.

## References

1. Rajkomar, A. et al. Scalable and accurate deep learning with electronic health records. npj Digital Medicine 1, 18 (2018).

2. Rajkomar, A., Dean, J. & Kohane, I. Machine Learning in Medicine. N. Engl. J. Med. 380, 1347–1358 (2019).

3. Obermeyer, Z. & Weinstein, J. N. Adoption of Artificial Intelligence and Machine Learning Is Increasing, but Irrational Exuberance Remains. NEJM Catalyst vol. 1 (2020).

4. Beam, A. L. & Kohane, I. S. Big Data and Machine Learning in Health Care. JAMA vol. 319 1317 (2018).

5. Goldstein, B. A., Navar, A. M., Pencina, M. J. & Ioannidis, J. P. A. Opportunities and challenges in developing risk prediction models with electronic health records data: a systematic review. J. Am. Med. Inform. Assoc. 24, 198–208 (2017).

6. Seneviratne, M. G., Shah, N. H. & Chu, L. Bridging the implementation gap of machine learning in healthcare. BMJ Innovations 6, 45–47 (2019).

7. Emanuel, E. J. & Wachter, R. M. Artificial Intelligence in Health Care: Will the Value Match the Hype? JAMA 321, 2281–2282 (2019).

8. Shah, N. H., Milstein, A. & Bagley, S. C. Making Machine Learning Models Clinically Useful. JAMA (2019) doi: 10.1001/jama.2019.10306.

9. Morse, K. E., Bagley, S. C. & Shah, N. H. Estimate the hidden deployment cost of predictive models to improve patient care. Nat. Med. 26, 18–19 (2020).

10. Sendak, M. P., Balu, S. & Schulman, K. A. Barriers to Achieving Economies of Scale in Analysis of EHR Data: A Cautionary Tale. Appl. Clin. Inform. 8, 826 (2017).

11. Sendak, M., Gao, M., Nichols, M., Lin, A. & Balu, S. Machine Learning in Health Care: A Critical Appraisal of Challenges and Opportunities. eGEMs 7, (2019).

12. Char, D. S., Shah, N. H. & Magnus, D. Implementing Machine Learning in Health Care — Addressing Ethical Challenges. N. Engl. J. Med. 378, 981 (2018).

13. Einav, L., Finkelstein, A., Mullainathan, S. & Obermeyer, Z. Predictive modeling of U.S. health care spending in late life. Science 360, 1462–1465 (2018).

14. Obermeyer, Z., Powers, B., Vogeli, C. & Mullainathan, S. Dissecting racial bias in an algorithm used to manage the health of populations. Science 366, 447–453 (2019).

15. Gade, G. et al. Impact of an Inpatient Palliative Care Team: A Randomized Controlled Trial. J. Palliat. Med. 11, 180–190 (2008).

16. Ma, J. et al. Early Palliative Care Consultation in the Medical ICU: A Cluster Randomized Crossover Trial. Crit. Care Med. (2019) doi: 10.1097/CCM.0000000000004016.

17. Smith, S., Brick, A., O’Hara, S. & Normand, C. Evidence on the cost and cost-effectiveness of palliative care: a literature review. Palliat. Med. 28, 130–150 (2014).

18. Temel, J. S. et al. Early palliative care for patients with metastatic non-small-cell lung cancer. N. Engl. J. Med. 363, 733–742 (2010).

19. Sullivan, D. R. et al. Association of Early Palliative Care Use With Survival and Place of Death Among Patients With Advanced Lung Cancer Receiving Care in the Veterans Health Administration. JAMA Oncol (2019) doi: 10.1001/jamaoncol.2019.3105.

20. Verret, D. & Rohloff, R. M. The value of palliative care. Healthc. Financ. Manage. 67, 50–54 (2013).

21. Osagiede, O. et al. Palliative Care Use Among Patients With Solid Cancer Tumors: A National Cancer Data Base Study. J. Palliat. Care 33, 149–158 (2018).

22. Evans, B. A., Turner, M. C., Gloria, J. N., Pickett, L. C. & Galanos, A. N. Palliative Care Consultation Is Underutilized in Critically Ill General Surgery Patients. Am. J. Hosp. Palliat. Care 37, 149–153 (2020).

23. Rubens, M. et al. Palliative Care Consultation Trends Among Hospitalized Patients With Advanced Cancer in the United States, 2005 to 2014. Am. J. Hosp. Palliat. Care 36, 294–301 (2019).

24. Wiskar, K., Toma, M. & Rush, B. Palliative care in heart failure. Trends Cardiovasc. Med. 28, 445–450 (2018).

25. Avati, A., Jung, K., Harman, S. & Downing, L. Improving palliative care with deep learning. (BIBM), 2017 IEEE … (2017).

26. Avati, A., Duan, T., Jung, K., Shah, N. H. & Ng, A. Countdown Regression: Sharp and Calibrated Survival Predictions. arXiv preprint 1806.08324 (2018).

27. Parikh, R. B. et al. Machine Learning Approaches to Predict 6-Month Mortality Among Patients With Cancer. JAMA Netw Open 2, e1915997 (2019).

28. Courtright, K. R. et al. Electronic Health Record Mortality Prediction Model for Targeted Palliative Care Among Hospitalized Medical Patients: a Pilot Quasi-experimental Study. J. Gen. Intern. Med. (2019) doi: 10.1007/s11606-019-05169-2.

29. Liu, V. X., Bates, D. W., Wiens, J. & Shah, N. H. The number needed to benefit: estimating the value of predictive analytics in healthcare. J. Am. Med. Inform. Assoc. 26, 1655–1659 (2019).

30. McIntosh, E., Donaldson, C. & Ryan, M. Recent advances in the methods of cost-benefit analysis in healthcare. Matching the art to the science. Pharmacoeconomics 15, 357–367 (1999).

31. Pantilat, S. Z., O’Riordan, D. L., Dibble, S. L. & Landefeld, C. S. Hospital-based palliative medicine consultation: a randomized controlled trial. Arch. Intern. Med. 170, 2038–2040 (2010).

32. Weiner, M. G., Sheikh, W. & Lehmann, H. P. Interactive Cost-benefit Analysis: Providing Real-World Financial Context to Predictive Analytics. AMIA Annu. Symp. Proc. 2018, 1076–1083 (2018).

33. Vickers, A. J. & Elkin, E. B. Decision curve analysis: a novel method for evaluating prediction models. Med. Decis. Making 26, 565–574 (2006).

34. Ke, G. et al. LightGBM: A Highly Efficient Gradient Boosting Decision Tree. in Advances in Neural Information Processing Systems 30 (eds. Guyon, I. et al.) 3146–3154 (Curran Associates, Inc., 2017).

35. Lowe, H. J., Ferris, T. A., Hernandez, P. M. & Weber, S. C. STRIDE--An integrated standards-based translational research informatics platform. AMIA Annu. Symp. Proc. 2009, 391–395 (2009).

36. Inflation Rate for Medical care between 2002-2019. https://www.in2013dollars.com/Medical-care/price-inflation/2002-to-2019?amount=1000.

37. Per person spending - Peterson-Kaiser Health System Tracker. Peterson-Kaiser Health System Tracker https://www.healthsystemtracker.org/indicator/spending/per-capita-spending/.

38. Scrucca, L. Model-based SIR for dimension reduction. Comput. Stat. Data Anal. 55, 3010–3026 (2011).

39. Shilo, S., Rossman, H. & Segal, E. Axes of a revolution: challenges and promises of big data in healthcare. Nat. Med. 26, 29–38 (2020).

40. Challener, D. W., Prokop, L. J. & Abu-Saleh, O. The Proliferation of Reports on Clinical Scoring Systems: Issues About Uptake and Clinical Utility. JAMA 321, 2405–2406 (2019).

41. Escobar, G. J. et al. Early detection of impending physiologic deterioration among patients who are not in intensive care: development of predictive models using data from an automated electronic medical record. J. Hosp. Med. 7, 388–395 (2012).

42. Escobar, G. J. & Dellinger, R. P. Early detection, prevention, and mitigation of critical illness outside intensive care settings. J. Hosp. Med. 11 Suppl 1, S5–S10 (2016).

43. Escobar, G. J. et al. Piloting electronic medical record-based early detection of inpatient deterioration in community hospitals. J. Hosp. Med. 11 Suppl 1, S18–S24 (2016).

44. Dummett, B. A. et al. Incorporating an Early Detection System Into Routine Clinical Practice in Two Community Hospitals. J. Hosp. Med. 11 Suppl 1, S25–S31 (2016).

45. Crown, W. et al. Constrained Optimization Methods in Health Services Research-An Introduction: Report 1 of the ISPOR Optimization Methods Emerging Good Practices Task Force. Value Health 20, 310–319 (2017).

46. Crown, W. et al. Application of Constrained Optimization Methods in Health Services Research: Report 2 of the ISPOR Optimization Methods Emerging Good Practices Task Force. Value Health 21, 1019–1028 (2018).

47. Beede, E. et al. A Human-Centered Evaluation of a Deep Learning System Deployed in Clinics for the Detection of Diabetic Retinopathy. in Proceedings of the 2020 CHI Conference on Human Factors in Computing Systems 1–12 (Association for Computing Machinery, 2020).

48. Li, R., Asch, S. & Shah, N. H. Developing a Delivery Science for Artificial Intelligence in Healthcare. npj Digital Medicine In press, (2020).

49. Beaulieu-Jones, B. et al. Trends and Focus of Machine Learning Applications for Health Research. JAMA Netw Open 2, e1914051 (2019).

50. Dalca, A. V. et al. Machine Learning for Health (ML4H) 2019 : What Makes Machine Learning in Medicine Different ? in Proceedings of the Machine Learning for Health NeurIPS Workshop (eds. Dalca, A. V. et al.) vol. 116 1–9 (PMLR, 2020).

